# Scoring policies with stakeholders to support healthy, sustainable food (kai) in Aotearoa New Zealand

**DOI:** 10.64898/2026.01.16.26344283

**Authors:** Bruce Kidd, Hemi Enright, Christina McKerchar, Christine Cleghorn

## Abstract

Engaging with stakeholders/contributors is vital to achieve a comprehensive approach to policymaking about the food (kai) system. The food system in Aotearoa New Zealand (Aotearoa NZ) is associated with high environmental impacts and inequitably distributed health consequences by ethnicity and socio-economic status. We engaged with contributors from government agencies, industry, academics, community, rural and urban Māori communities to develop policy actions to support New Zealanders to eat healthy sustainable food. This paper outlines methods used to score contributor identified policies with follow-up focus groups and interviews where the highest scored policies were discussed, to understand contributor perspectives on the scope, implementation, barriers, and possible unintended consequences of these policies. Semi-structured interviews (12) and focus groups (6) were conducted. The top five scored policies among all contributors were: healthy food and drink policies in schools and kura (Māori-language immersion schools); supporting māra kai (food garden) and community gardens; garden to table programmes in schools and kura; education about sustainable and healthy food; and increasing incomes. The top five policies for Māori were: healthy food and drink policies in schools and kura; garden to table programmes in schools and kura; remove GST from core foods; increase incomes and make local food cheaper by supporting local growers. Considerations for policy implementation involved implementing multiple policies at once, the importance of evaluation, finding existing examples of policies, strong collaboration for long-term outcomes, and centring policies through a Māori framework.

## Introduction

Enabling dietary transitions to healthy and sustainable kai (food) can reduce health and environmental impacts resulting from our global food system [1–4]. Currently, global food systems contribute 34% of total greenhouse gas (GHG) emissions from activities relating to agriculture and land use, storage, transport, packaging, processing and food retail [5]. Environmental effects include but are not limited to, loss of habitat and biodiversity of native species and deterioration of water resources. In 2023 the agricultural industry in Aotearoa New Zealand (NZ) accounted for 53% of gross GHG emissions [6].

Dietary risks and high body mass index were the second and third highest contribution to health loss as measured by Disability Adjusted Life Years (DALYs) in Aotearoa NZ [7]. Diet-related health outcomes such as obesity are inequitably distributed due to a failure of policy. In relation to Māori the Indigenous people of NZ these failures represent a breach of Te Tiriti o Waitangi (a treaty signed in 1840 by the Crown and Māori) that includes the right to health equity [8,9]. Focusing on food systems can produce planetary health gains such as reducing GHG emissions and therefore alleviate the consequences of climate change on food security.

To promote planetary health, the EAT-Lancet Commission [3,4] since 2019 has proposed an internationally acceptable reference diet (“planetary diet”) based on Rockström et al.’s [10] planetary boundaries. The planetary diet depicts a sustainable and healthy diet as high in unprocessed plant-based foods and low in animal-origin foods, consistent with the international literature [11–13]. In 2025 the planetary diet was updated by reviewing the social foundations of a just food system, depicting how plant-rich and culturally adaptable diets can benefit people [4]. Utilising a food systems approach to policy development and implementation requires strong engagement with affected stakeholders and communities [14–16]. It is important to investigate what mix of policies are likely to be adopted by investigating the feasibility, acceptability and health equity impacts of any intervention.

Research indicates that strong engagement with stakeholders and policy-level support, involving collaboration at different levels such as national and local, are essential enablers [17–20]. In the first stage of this research project contributors, a term to describe our participants inspired by Māori kupu (words) like kairaranga (weaver), provided policy suggestions on how to support New Zealanders to consume healthy and sustainable kai. Taking this collective approach to policymaking is important when considering food system policies.

This current paper follows on from our previous publication outlining the first stage of the research [21], detailing the policy scoring exercise for identified policies (a variation of the Assessing Cost-Effectiveness (ACE) approach to priority setting) and follow-up interviews and focus groups. These interviews and focus groups aimed to define the policies and outline their scope, implementation, barriers and possible unintended consequences.

## Methods

The policy scoring exercise informed part of a wider project. A qualitative process first identified policies which then contributed to quantitative modelling of health impacts [22]. The 111 policies identified through the initial focus groups and interviews were rapidly reviewed to ensure sufficient evidence of effectiveness to be modelled through to health gains, methods used in the subsequent stage of the project [23,24]. Those policies with sufficient evidence were included in an online scoring exercise emailed to contributors involved in the initial consultation and project advisors. The six policies that scored highest were taken through to the final stage of consultation, which discussed how these six policies should be implemented in Aotearoa NZ.

### Rapid review

Two public health nutrition databases (Scopus; Nutrition and Food Sciences Database) were used to determine if the suggested policies had sufficient high-quality evidence available on their effectiveness in changing dietary intake. Each policy’s inclusion criteria and search terms varied depending on the policy and the breadth of literature currently available. For example, if there was known Aotearoa NZ-based literature for a selected policy, this was stated in the inclusion criteria. If no Aotearoa NZ-based literature was available, international evidence was substituted in the inclusion criteria. This process resulted in groupings of similar policies.

After noting the availability and quality of evidence, the initial round of exclusion involved assessing whether the policy could have an impact on dietary risk factors (e.g. fruit and vegetable intake) and if the scope was not too broad or the impact too distant from consumption to model. Then each policy was assessed on its plausibility to be modelled. It was labelled as “yes,” “no,” or “unsure”. All policies deemed “unsure” or “yes,” had their relevant literature assessed further before inclusion. Policies considered as “no,” were noted as unlikely to be modelled and excluded. The principal investigator completed the determination of modelling plausibility on this project. Dr Cristina Cleghorn (CC) has extensive experience and expertise in modelling policy and intervention effectiveness and the cost-effectiveness of dietary interventions in Aotearoa NZ’s population [23,24].

Of the 111 unique policies identified in the initial consultation, sixty-three were excluded as there was no direct pathway to impact on dietary consumption or they were too broad to be modelled in the Multistate Life Table (MSLT) model. The remaining 48 policy suggestions were rapidly reviewed, resulting in 14 policies with evidence that could be scored in the project’s next stage.

### Policy scoring

A variation of the ACE approach to priority setting was applied before instigating cost effectiveness analyses. The ACE approach is summarised in Figure 1 of the paper Carter et al. (2008) [25] on setting priorities in health. The ACE research group in Australia has successfully applied this method to prioritise 150 preventive health interventions [25,26]. The ACE research group used a broad decision-making framework that included considerations of equity, strength of evidence, feasibility of implementation, contributor acceptability, and other study-specific considerations [25,26]. These were used alongside the cost-effectiveness results for policy-makers to judge the interventions. The ACE report details methods that were adapted for this project [27].

**Figure 1:**
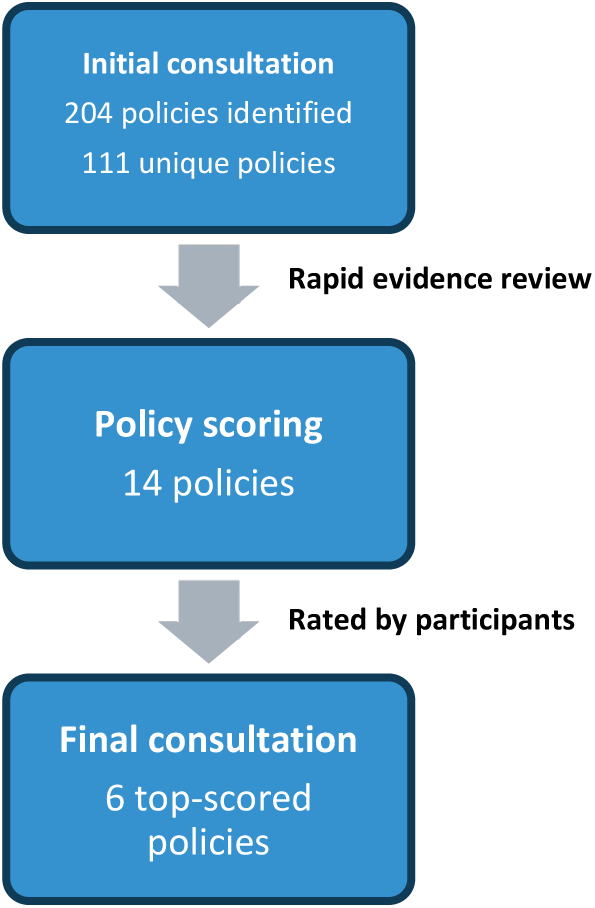
Overview of the policy selection process.

Policies were scored based on the following five criteria (see Supplementary Supplementary Table 3 for details): the acceptability and feasibility of the policy, ability to improve the environment; diets of New Zealanders; and Māori health. These criteria were used to score policies on a Likert scale from one to five. For example, we asked contributors, “please enter your rating on the capacity for [policy X] to [criterion Y]” on the following scale: one – Poor; two – Fair; three – Good; four – Very good; five – Excellent; and Cannot rate. All the fields in the scoring exercise were required to be completed by the respondent, except the further comments box.

The scoring exercise was piloted among the project team and staff members at the University of Otago, particularly the Public Health department to ensure the criteria and the process would be clear to contributors. Feedback was incorporated in the final version. The scoring exercise was emailed to contributors and collected information on age group, gender, ethnicity, and contributor group. For groups that preferred group interaction or requested additional support for the scoring exercise, an online Zoom meeting was provided.

### Existing contributors

Previous focus groups were held with individuals from government ministries, community members and contributors from two Māori communities (urban and rural). Interviews were also completed with academics and industry, which included farmers. These interviewees were defined as existing contributors. All individuals involved in focus groups or interviews were given the opportunity to participate in the scoring stage of the consultation as well as the project’s advisory group. A supplementary document (Supplementary Supplementary Table 3) was provided to individuals, which summarised each policy mentioned in the exercise and defined each criterion used to assess the policy.

All contributors were given three weeks to complete the scoring exercise, with reminder emails sent at weeks one and two. REDCap was used to enable completion tracking. An ethics amendment was submitted on 12 October 2022 and approved on 13 October 2022 to enable the “participant identifier” feature in REDCap. When there was no email address for a focus group contributor, we sent a live link to the focus group co-ordinator to disseminate to attendees.

Once scoring exercise responses were completed, data were exported from REDCap into a .csv format compatible with Microsoft Excel. A criterion completed with “cannot rate” was given the average score of the ratings provided for the same criterion from the remaining contributors. This method was used as we assumed contributors chose this option as they felt they had insufficient expertise to answer. A total score and mean score were calculated across all five criterions for each policy. These scores were calculated separately for “Māori contributors only” and for “all contributors (including Māori)” to ensure the perspectives of Māori contributors were distinct. The policies that were in the top five among “all contributors” or among “Māori contributors” were compared and progressed through to the next stage of the consultation. When presenting the data we converted the numerical variable of the score (1-5) to a categorical variable to convey the key points from the data. Scores one to two were grouped as “disagree”, score three as “neutral”, and scores four and five as “agree”. Once the analysis was finalised and the selected policies progressing to the next stage were determined, the six top-scored policies were discussed with contributors in focus groups and interviews.

### Focus groups and interviews

All original contributors were invited to take part in focus groups and semi-structured interviews to discuss the six top-scored policies. Researcher questions in the focus groups and interviews focused on the policies’ scope, implementation, barriers and how these barriers could be overcome (specifically concerning policy design), and potential unintended consequences. At the beginning of each interview or focus group, the research team summarised key results, aims and objectives of the session. Policy specific prompt questions were asked when points had not already been covered, for example:

- How do you think this should be implemented in NZ?
- How could the policy work at a national level?
- Who is likely to be involved in this project and who could benefit from it?
- What resources will need to be put in place for this to work?

We asked contributors for their views on each six top-scored policies and whether they agreed or disagreed with the policy being modelled in the next stage of the project (Figure 1).

Focus groups and interviews were recorded with contributor consent and transcribed using the software “Otter.AI.” Public health research is increasingly using Artificial Intelligence (AI) to streamline tasks and reduce human workload [28]. We acknowledge that AI has predominantly been developed by non-Māori without consideration of the health context in Aotearoa, Kaupapa Māori ethics or Māori Data Sovereignty, which recognise that Māori data should be subject to Māori governance [29–31]. However, given the utility of Otter.AI which allowed us to create an initial draft of transcripts and playback audio files while making amendments to the text, we chose to use it in this context but ensured that a Māori researcher replayed and checked (line by line) each transcription where Māori contributors were interviewed. The research team reflected on the use of Otter.AI during the research process and since completion, removed all audio files and transcriptions from the platform.

Descriptive coding was used as part of a pragmatic approach to our mixed methods project to assess transcripts for the key themes of scope, considerations for implementation, barriers, and possible unintended consequences among other criteria. Using thematic analysis, transcripts were reviewed, and information was collated in Microsoft Excel. The transcription review process ensured that all information was captured to further define policies. Themes were then generated for each policy. These themes were developed either when more than one contributor expressed a similar point or from a single mention if there was little information on the policy during the discussion. Several contributors used Māori words and these are defined in the glossary section. In this paper, the kupu “kura” refers to Māori-language immersion schools.

## Results

Fourteen policies with evidence of effectiveness were selected (Table 1) to undergo policy scoring. Most of these policies were from the food environment policy domain. The rating exercise was sent to fifty-nine contributors from the initial consultation and advisory group. Thirty-seven full responses (every policy rated) and five partial responses (at least one policy rated) were received (we had no response from seventeen), with an overall 71% response rate. Twelve full and one partial response were from Māori. In the next stage, all focus group contributors and interviewees were retained except for contributors who were no longer available i.e. had left their organisation following initial consultation. This resulted in an average of five contributors (twenty-eight total) in each focus group and twelve individual interviews to discuss the highest ranked policies.

**Table 1:**
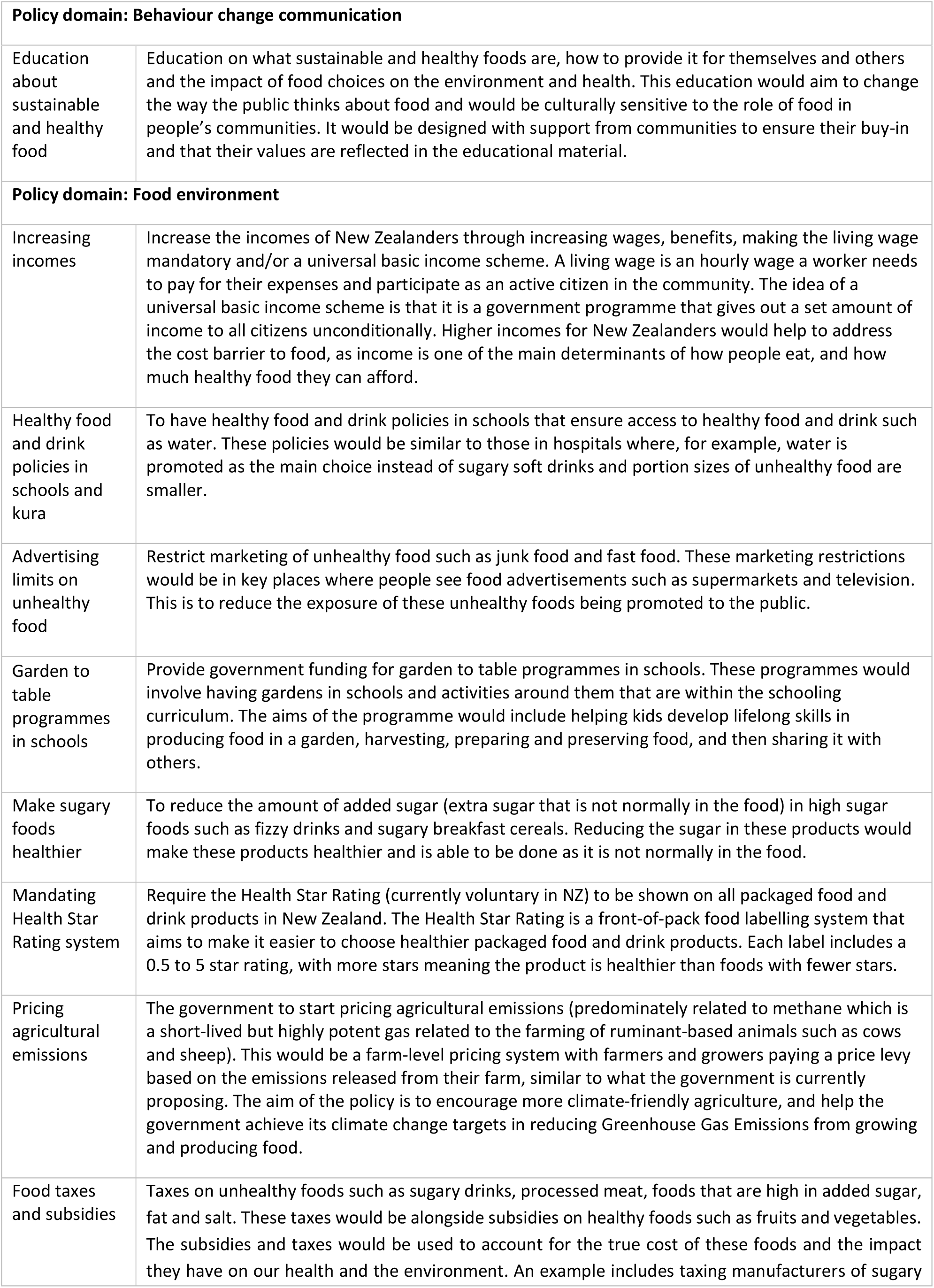
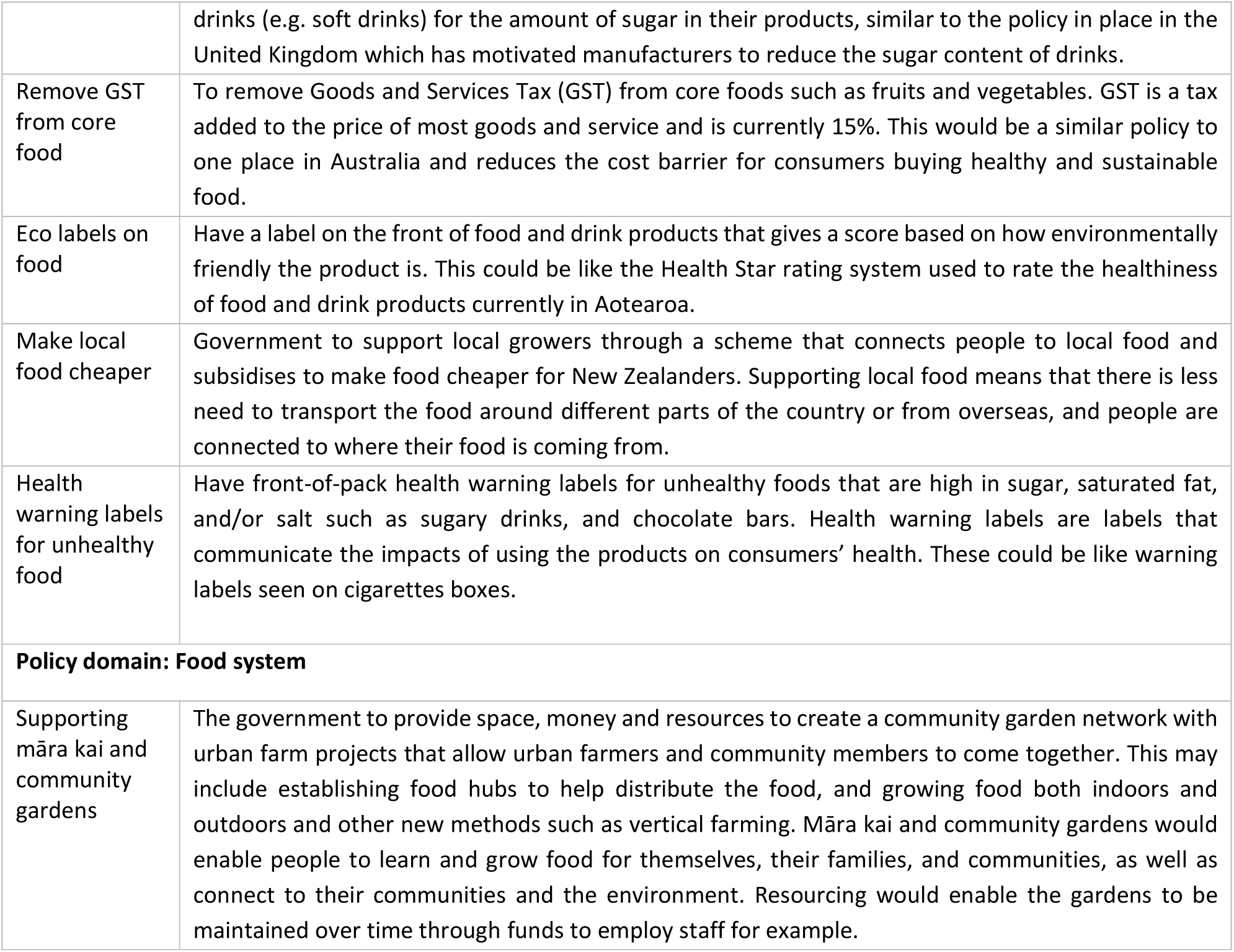
Overview of policies selected for the policy scoring exercise by NOURISHING framework policy domains [32].

|  |
| --- |
| Policy domain: Behaviour change communication |

|  |  |
| --- | --- |
| Education about sustainable and healthy food | Education on what sustainable and healthy foods are, how to provide it for themselves and others and the impact of food choices on the environment and health. This education would aim to change the way the public thinks about food and would be culturally sensitive to the role of food in people's communities. It would be designed with support from communities to ensure their buy-in and that their values are reflected in the educational material. |
| **Policy domain: Food environment** |  |
| Increasing incomes | Increase the incomes of New Zealanders through increasing wages, benefits, making the living wage mandatory and/or a universal basic income scheme. A living wage is an hourly wage a worker needs to pay for their expenses and participate as an active citizen in the community. The idea of a universal basic income scheme is that it is a government programme that gives out a set amount of income to all citizens unconditionally. Higher incomes for New Zealanders would help to address the cost barrier to food, as income is one of the main determinants of how people eat, and how much healthy food they can afford. |
| Healthy food and drink policies in schools and kura | To have healthy food and drink policies in schools that ensure access to healthy food and drink such as water. These policies would be similar to those in hospitals where, for example, water is promoted as the main choice instead of sugary soft drinks and portion sizes of unhealthy food are smaller. |
| Advertising limits on unhealthy food | Restrict marketing of unhealthy food such as junk food and fast food. These marketing restrictions would be in key places where people see food advertisements such as supermarkets and television. This is to reduce the exposure of these unhealthy foods being promoted to the public. |
| Garden to table programmes in schools | Provide government funding for garden to table programmes in schools. These programmes would involve having gardens in schools and activities around them that are within the schooling curriculum. The aims of the programme would include helping kids develop lifelong skills in producing food in a garden, harvesting, preparing and preserving food, and then sharing it with others. |
| Make sugary foods healthier | To reduce the amount of added sugar (extra sugar that is not normally in the food) in high sugar foods such as fizzy drinks and sugary breakfast cereals. Reducing the sugar in these products would make these products healthier and is able to be done as it is not normally in the food. |
| Mandating Health Star Rating system | Require the Health Star Rating (currently voluntary in NZ) to be shown on all packaged food and drink products in New Zealand. The Health Star Rating is a front-of-pack food labelling system that aims to make it easier to choose healthier packaged food and drink products. Each label includes a 0.5 to 5 star rating, with more stars meaning the product is healthier than foods with fewer stars. |
| Pricing agricultural emissions | The government to start pricing agricultural emissions (predominately related to methane which is a short-lived but highly potent gas related to the farming of ruminant-based animals such as cows and sheep). This would be a farm-level pricing system with farmers and growers paying a price levy based on the emissions released from their farm, similar to what the government is currently proposing. The aim of the policy is to encourage more climate-friendly agriculture, and help the government achieve its climate change targets in reducing Greenhouse Gas Emissions from growing and producing food. |
| Food taxes and subsidies | Taxes on unhealthy foods such as sugary drinks, processed meat, foods that are high in added sugar, fat and salt. These taxes would be alongside subsidies on healthy foods such as fruits and vegetables. The subsidies and taxes would be used to account for the true cost of these foods and the impact they have on our health and the environment. An example includes taxing manufacturers of sugary |
|  | drinks (e.g. soft drinks) for the amount of sugar in their products, similar to the policy in place in the United Kingdom which has motivated manufacturers to reduce the sugar content of drinks. |
| Remove GST from core food | To remove Goods and Services Tax (GST) from core foods such as fruits and vegetables. GST is a tax added to the price of most goods and service and is currently 15%. This would be a similar policy to one place in Australia and reduces the cost barrier for consumers buying healthy and sustainable food. |
| Eco labels on food | Have a label on the front of food and drink products that gives a score based on how environmentally friendly the product is. This could be like the Health Star rating system used to rate the healthiness of food and drink products currently in Aotearoa. |
| Make local food cheaper | Government to support local growers through a scheme that connects people to local food and subsidises to make food cheaper for New Zealanders. Supporting local food means that there is less need to transport the food around different parts of the country or from overseas, and people are connected to where their food is coming from. |
| Health warning labels for unhealthy food | Have front-of-pack health warning labels for unhealthy foods that are high in sugar, saturated fat, and/or salt such as sugary drinks, and chocolate bars. Health warning labels are labels that communicate the impacts of using the products on consumers' health. These could be like warning labels seen on cigarettes boxes. |
| **Policy domain: Food system** |  |
| Supporting māra kai and community gardens | The government to provide space, money and resources to create a community garden network with urban farm projects that allow urban farmers and community members to come together. This may include establishing food hubs to help distribute the food, and growing food both indoors and outdoors and other new methods such as vertical farming. Māra kai and community gardens would enable people to learn and grow food for themselves, their families, and communities, as well as connect to their communities and the environment. Resourcing would enable the gardens to be maintained over time through funds to employ staff for example. |

### Scoring exercise

The top scoring policies from the scoring exercise were analysed for overall respondents and for Māori alone. In descending order, the top five scoring policies among all contributors were: healthy food and drink policies in schools and kura; supporting māra kai and community gardens; garden to table programmes in schools and kura; education about sustainable and healthy food; and increasing incomes. For Māori the top five scoring policies were: healthy food and drink policies in schools and kura; garden to table programmes in schools and kura; remove GST (Goods and Services Tax) from core food; increasing incomes and make local food cheaper.

Contributors’ level of agreement varied by policy and criteria (Table 2, Figure 2). The policies with the highest agreed acceptability were supporting māra kai and community gardens (93%), education about sustainable and healthy food (90%), garden to table programmes in schools and kura (86%), and healthy food and drink policies in schools and kura (83%). A large proportion agreed that increasing incomes (90%), healthy food and drink policies in schools and kura (88%), removing GST from core food (88%), food taxes and subsidies (81%), and making local food cheaper (71%) would improve Māori health.

**Figure 2:**
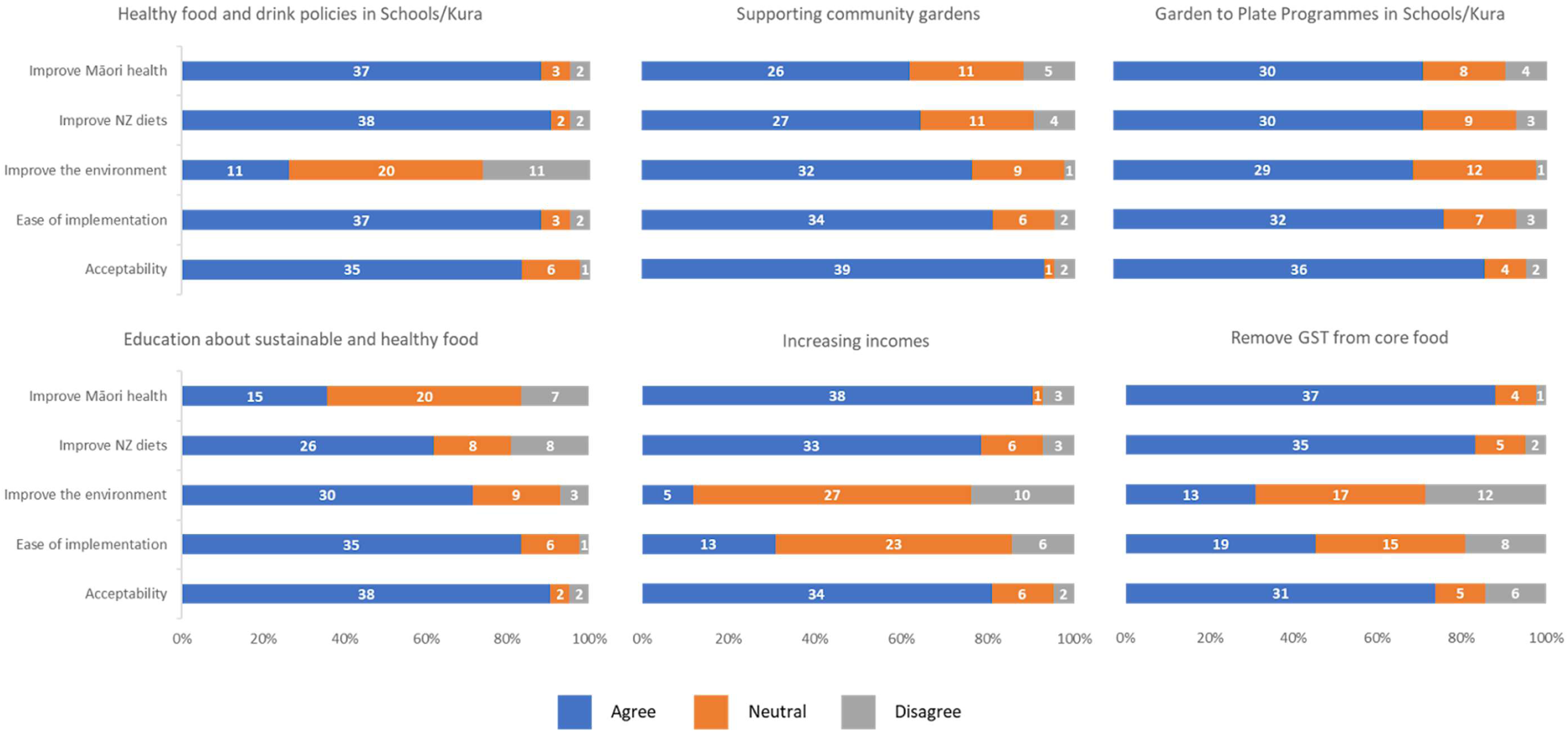
Distribution of agreement on top six policies according to policy scoring criteria.

**Table 2:** Overall rank and scores for policies for all participants and Māori participants according to the criteria.

| Policy | Overall ranking |  | Improve Māori health |  | Improve NZ Diets |  | Improve the environment |  | Ease of implementation |  | Acceptability |  |
| --- | --- | --- | --- | --- | --- | --- | --- | --- | --- | --- | --- | --- |
|  | All* | Māori ** | All | Māori | All | Māori | All | Māori | All | Māori | All | Māori |
| Healthy food and drink policies in schools and kura | 1 | 1 | 4.2 | 4.5 | 4.3 | 4.3 | 3.2 | 4.5 | 4.3 | 4.5 | 4.2 | 4.5 |
| Supporting māra kai and community gardens | 2 | 6 | 4.4 | 4.8 | 4.1 | 3.9 | 4 | 4.8 | 3.8 | 4.8 | 3.7 | 4.8 |
| Garden to table programmes in schools and kura | 3 | 2 | 4.2 | 4.5 | 4 | 4.3 | 3.8 | 4.5 | 4 | 4.5 | 3.8 | 4.5 |
| Education about sustainable and healthy food | 4 | 7 | 4.3 | 4.4 | 4.2 | 4 | 3.8 | 4.4 | 3.6 | 4.4 | 3.4 | 4.4 |
| Increasing incomes | 5 | 4 | 4.1 | 4.4 | 3.4 | 4.3 | 2.9 | 4.4 | 4.1 | 4.4 | 4.5 | 4.4 |
| Remove GST from core food | 6 | 3 | 3.9 | 4.7 | 3.4 | 4.4 | 3.1 | 4.7 | 4.1 | 4.7 | 4.2 | 4.7 |
| Make local food cheaper | 7 | 5 | 4.1 | 4.6 | 3.2 | 4 | 3.7 | 4.6 | 3.8 | 4.6 | 4 | 4.6 |
| Advertising limits on unhealthy food | 8 | 8 | 3.8 | 4.2 | 3.9 | 4.1 | 2.8 | 4.2 | 3.9 | 4.2 | 3.6 | 4.2 |
| Food taxes and subsidies | 9 | 9 | 3.1 | 3.5 | 2.9 | 3.8 | 3.5 | 3.5 | 3.9 | 3.5 | 4.1 | 3.5 |
| Mandating a Health Star Rating System | 10 | 11 | 3.8 | 4.4 | 3.5 | 3.8 | 2.3 | 4.4 | 3.3 | 4.4 | 3.1 | 4.4 |
| Eco labels on food | 11 | 10 | 3.6 | 4.3 | 3.2 | 3.8 | 3.5 | 4.3 | 2.7 | 4.3 | 2.8 | 4.3 |
| Make sugary foods healthier | 12 | 12 | 3.5 | 3.8 | 2.8 | 3.4 | 2.1 | 3.8 | 3.6 | 3.8 | 3.6 | 3.8 |
| Health warning labels for unhealthy food | 13 | 13 | 3.3 | 3.9 | 2.9 | 3.7 | 2.3 | 3.9 | 3.4 | 3.9 | 3.2 | 3.9 |
| Pricing agricultural emissions | 14 | 14 | 2.9 | 3.5 | 2.8 | 2.9 | 4 | 3.5 | 2.6 | 3.5 | 2.9 | 3.5 |

Most contributors agreed that healthy food and drink policies in schools and kura (88%), education about sustainable and healthy food (83%), supporting community gardens (81%), and garden to table programmes in schools and kura (76%) were easier to implement. The majority considered that healthy food and drink policies in schools and kura (90%), removing GST from core food (83%), increasing incomes (79%), food taxes and subsidies (76%), and advertising limits on unhealthy food policies (76%) would improve NZ diets. The highest proportion of respondents agreed that the following policies: pricing agricultural emissions (74%), supporting community gardens (76%) and education about sustainable and healthy food (71%) would improve the environment.

Due to the overlapping evidence to be used in modelling between the ‘increasing incomes’ and ‘make local food cheaper’ policy, a decision was reached to select only one of the two policies to take forward into stage three. The latter policy was not progressed as ’increasing incomes’ received a higher ranking and overall score from Māori contributors and all contributors. The six policies included in the top five policies in either all participants or Māori alone were: supporting māra kai and community gardens; garden to table programmes in schools and kura; healthy food and drink policies in schools and kura; education about sustainable and healthy food; removing GST from core food; and increasing incomes. Table 2 identifies the full scores and rankings for policies. Figure 2 shows the distribution of agreement between contributors on key policies.

### Focus group and interviews

The top six scored policies were discussed in more detail during focus groups and interviews. Contributors spoke of general considerations for policy implementation, barriers to policies and unintended consequences. Essential considerations for implementation involved implementing multiple policies at once, the importance of evaluation, finding existing examples of policies and building on these, strong collaboration for long-term outcomes, and centring the policies through a Māori framework. Grouping the policies was suggested as part of a policy package for the greatest impact. One contributor spoke of establishing a Ministry of Food to provide a home for all the policies to be designed, implemented and evaluated.

Many contributors recommended that policies be piloted before implementation. However, models for piloting were discussed as being too small to measure the policy’s benefits, especially concerning health outcomes that may take longer to appear. Additionally, designing and running a pilot may not recognise the existing resources in the community, particularly when this is done by an external agency. Concerns were raised about pilots, especially how the piloting process often fails to acknowledge and develop relationships with communities before commencement. Some contributors instead suggested funding community champions who have the knowledge, skills and resources to ensure long-term action. As elaborated by a contributor,

> *“… the trouble with piloting is that you set up expectations… the money runs out, all the supports and the resources that went with it disappear, and then you have actually done real harm to communities”* (urban Māori focus group).

Numerous contributors spoke of policies currently being developed in a siloed approach instead of building on existing community-driven initiatives. Contributors felt that by using what was already established, the need for a pilot was removed or reduced. Further to this, often evaluations of these projects identified the necessary development and implementation considerations needed to develop healthy public policy. Contributors spoke to a need for agencies to collaborate and focus on intergenerational outcomes when developing policy and initiatives for communities. Māori contributors raised the need for the policies to be centred through a Māori framework to *“reimagine”* (urban Māori focus group) policies. This included focusing on restoring the connections between land, people and food, to decolonise food intake and the overall food system. As one contributor elaborated,

> *“Because you see, this is presented to us as an English document (the provided prompts). Yep. But actually, we see it through a Māori framework the whole time. And that’s, you know, possibly, one of the ways to go would be to reimagine what those six things would look like from a framework that we might employ”* (rural Māori focus group).

### Healthy food and drink policies in schools and kura

Policy scope involved implementation across both primary and secondary schools. Contributors recognised that developing a healthy food environment in secondary schools or kura was imperative. Consensus was that the policy should apply only to food and drink sold in schools, excluding food and drinks bought from elsewhere. Key themes that emerged were the importance of community ownership and buy-in, promoting plant-based options as part of nutrition guidelines, integrating culture into policy, ensuring healthier food that aligns with policy is accessible, and packaging this with other initiatives such as school lunch programmes. Ensuring that the food in the policy is accessible was discussed by contributors such as making foods that fit under the policy cheaper and ensuring schools are well funded to not rely on food sales from unhealthy junk food.

Contributors spoke to how a bottom-up approach where the views and ideas of communities were centred was necessary for success. For kura, it was noted that many have had healthy kai policies for years, and there is a need to acknowledge this existing work. Integrating Māori, Pacific and kura specific culture into the policy was discussed as vital when defining what is healthy food to people and communities. A contributor provided an example of an unintended consequence when the views of community members were not acknowledged and integrated into policy, when they spoke about kale and how it may be recognised by policy writers as healthy kai but because it is not applicable to the child and their whānau context, it is not of use or used,

> *“I think we should have some beautiful kale because that’s nutritious. It’s got lots of vitamin C […], you know, what the hell’s this kale? It’s what do I do with that [kale]? And it will go in the bin. So there’s got to be a relationship to familiar kai (foods) as well.”* (urban Māori focus group)

Contributors identified that if a national approach to food in schools and kura were to be adopted this could lead to omission of mātauranga (Māori knowledge) and tikanga Māori (Māori custom, rules, practices and guidance) which would be a significant barrier to relevance and implementation in NZ kura and schools. One unintended consequence was burnout among participating schools to comply with the policy as part of not having the necessary resources (both human and monetary). Māori contributors also stated capacity within kura (in particular) is limited by both human and financial resources.

### Supporting māra kai and community gardens

Robust funding and centring community champions to lead initiatives were two central themes in the supporting māra kai and community gardens policy discussions. Māra are different to community gardens, a māra is a space where Rongo (atua of peace) is omnipotent and tangata whenua are connected to the whenua by story-telling intrinsic to the cultivation and consumption of kai borne of Māori whenua [33–36]. Contributors talked about Māra in this context.

Financial support is important for māra kai and community gardens, and contributors identified ongoing funding needs for seeds and bulk seedlings, garden maintenance, purchase of land, equipment for supplies and labour, and long-term roles associated with maintaining the space. Contributors agreed that funding should come from central government and be distributed to locally controlled and operated organisations. Ideally, there would be flexible, non-onerous criteria and processes when applying for funding that acknowledged the important work that is done and the commitment of these organisations to their people. Funded paid roles for community champions were highlighted as a critical requirement for long-term success of local initiatives and preventing burnout among staff. For māra kai, specific aspects were discussed such as using existing māra, as one contributor said,

> “*[…] all these gardens do really well because they’ve got a champion that’s true of all of them, and as soon as you pull that person out, the thing just dips and dies and fades”* (rural Māori focus group).

Other key supports needed were: high-value land being allocated preferentially, with areas classified as ’food deserts’ (areas with poor access to affordable and healthy food) prioritised first and the food hubs incorporated into the gardens as discussed in the initial consultation. Contributors spoke of barriers to implementation, such as gardens not being maintained due to a lack of long-term investment to support champions as well as maintaining the soil and grounds. Many contributors raised concerns about inequitable access with wealthier people more likely to be able to access gardens due to the time and resources people need to participate in a garden. For Māori, access to māra for urban marae was also discussed. An unintended consequence of the policy was the potential for harm to the land if a garden is not sustained, as the garden can become overgrown and hazardous.

### Garden to table programmes in schools and kura

Contributors identified key aspects of the policy as programme availability in all schools (primary and secondary), and the policy being encouraged instead of compulsory for these schools. There was a suggestion that the mandatory nature of the policy may make it difficult for all schools to comply,

> *“I don’t think it should apply to all schools, because not all schools will have the resources and ability to [comply]. I think it should be encouraged in all schools and applied where it makes sense”* (Farmer).

The main themes that emerged for this policy included connecting and expanding existing garden to table programmes, strong support for teachers and school staff, providing extra resourcing to integrate into the schooling curriculum, prioritising implementation in lower socioeconomic schools first, enabling local engagement with the gardens, and maintaining a roster over the holidays to ensure the gardens are maintained. Contributors also discussed implementing the policy alongside supporting māra kai and community gardens, free school meals, and healthy food and drink policies in schools to create a healthy space not only in the school but also the wider community so that children and their whānau could both benefit and make long-term changes to their consumption and cultivation of food.

Contributors highlighted the need for specific consideration for kura, including funding to support kura and their staff and a champion to push the kaupapa. Whānau involvement and engagement were also seen as essential for the programmes continued success in the community. Barriers to the programme’s success were thought to be: the capacity of school staff and availability of resources to engage with the programme, equity concerns from the cost associated with participation, security issues, access to quality land at school, and the ability to maintain the gardens over summer and school holidays.

### Education about sustainable and healthy food

The importance of implementing education alongside other policies for effectiveness, cross sectoral collaboration, building on existing education material and resources, utilising strong positive messaging with a primarily health focus was discussed. If implemented by itself, the policy may be ineffective and require other policies to support education. Contributors suggested that education should focus on key topics such as how Aotearoa NZ grows food, how food production affects people and the environment, promoting meat-free meals (at least once a week), being a food citizen, the use of critical thinking about food and broader food systems education. Contributors also highlighted the importance of having a separate framework for Māori,

> *“[…] I would hate to think that mainstream decides what’s a good, hauora (health) approach for Māori […] It can’t be done as just the kai policy or tīkanga. It has to be done so that it fits with other parts of what your te taha wairua (the spiritual health), te taha hinengāro (the mental and emotional health), whānau (social health and connection to others)”* (urban Māori focus group)

Contributors raised using existing education examples in the community, engaging with communities to review educational materials and ensuring central government provided funds to organisations working on healthy food education. A decolonising approach to dietary intake was discussed to ensure a holistic and comprehensive view as to what is “healthy” for Māori. Co-design and co-governance with Māori would need to occur throughout this policy.

What is defined as ‘healthy and sustainable kai’ was identified by contributors as being contentious, given that there is no agreed definition or government position on healthy and sustainable food in NZ. Equity concerns were also highlighted as education initiatives can worsen health equity as they are generally more effective for educated populations with higher literacy and more resources. Education interventions do not address issues such as the cost of healthy, sustainable food which impact lower income communities more. Additionally, contributors identified that a barrier to the policy may be strong commercial opposition, especially if the education promotes more plant-based than animal-based food products.

### Increasing incomes

Contributors expressed different views on the approaches to implementing this policy but were generally positive about the need to increase incomes, especially for lower income groups. The two scenarios were a mandatory living wage and a Universal Basic Income (UBI). A living wage is defined as the necessary income to provide the basic necessities of life for workers and their families, and enable their dignity and active participation in society. A UBI involves an unconditional payment to citizens with no conditions attached e.g. no paid work requirements. An alternative to UBI that was targeted to low-income people was discussed such as adding credit redeemable at community markets to a Community Services Card (used by low-income people in NZ to access goods and services at a subsidised cost).

The final scenario provided to contributors to discuss was fruit and vegetable vouchers as an alternative to a UBI. One contributor explained their support of both measures, as well as any measure designed to increase financial support to low income people that enable participation in the food system with the following quote;

> *“I think that having a mandatory living wage and a UBI […] could be really effective ideas. […] for example, […] allow some people to say, great, I can afford to work for four days and […] give a day to volunteering in the community garden”* (academic)

Contributors identified that the UBI was non-stigmatising, as it is for everyone and because of this it likely would have a low administrative requirement during implementation compared to policies targeted to specific groups. Therefore, contributors justified the UBI as being supportive of equitable access to healthy and sustainable food as by its very nature, the UBI may reduce the concentration of wealth among the wealthy and disproportionately benefit lower socio-economic individuals who may lack a regular, guaranteed income source.

If a living wage was implemented, the ability of businesses to pay these higher wages was discussed as a potential barrier and may result in opposition. The policy was also raised as being vulnerable to changes in government, with further instability from a short, three year electoral term in Aotearoa NZ. There was some disagreement between contributors that the UBI was not targeted enough to those who need it most. Unintended consequences included the general cost of living rising with higher wages and money not being spent on healthy food.

### Remove Goods and Services Tax (GST) from core food

Versions of this policy were suggested by different contributors, most were generally supportive of removing GST if it led to reduced food prices. Some contributors suggested focusing on the core food definition associated with the existing Australian policy, adding sustainability to the ’core’ food definition, focusing on just fruits and vegetables, or just local fruits and vegetables [37]. Contributors spoke of learning from Australia, ensuring more significant benefit of the policy for those on lower incomes, including strong engagement with retailers and providing recipe ideas for foods with GST removed. Several key barriers were identified by contributors such as loss of government revenue from not collecting GST on core foods, the difficulty associated with defining what constitutes a core food and what is a sustainable food as well as administration costs with implementing the policy within the current taxation system.

Several unintended consequences and additional considerations were raised by contributors. Contributors discussed the likelihood of the meat and dairy industry lobbying against the policy if meat and dairy were not defined as core foods and therefore eligible for GST exemption. Other possible unintended consequences included retailers absorbing the price decrease and not passing on these savings to customers. Contributors spoke of the nature of the tax system in Aotearoa NZ, being relatively straightforward in how GST is administered. By introducing an exemption for selected foods this may create additional administration costs. The loss of government funding from GST may result in health initiatives being negatively impacted. One contributor spoke to how this policy could increase inequities,

> “*I think that is an ….. inequity increasing policy. Because I think the people who would benefit most from that are us. And that would be a loss of revenue to the government that could actually have been targeted to supporting those who are less able to afford fruits and vegetables”* (Academic).

Another equity consideration was that wealthier individuals (as measured by income) have more money to spend on food so will receive greater savings. Wealthier individuals can then buy more fresh fruit and vegetables and benefit from this, increasing the gap in health outcomes between higher income whānau and lower income groups.

## Discussion

Most policies included in the scoring survey focused on food environments which is not surprising given that food environment policies and targeted interventions are more likely to be extensively researched. In contrast, food system domain policies were often too broad to have specific evidence of effect on dietary consumption, reflected in their lower rates of inclusion. The top policies from the scoring exercise were supporting māra kai and community gardens, garden to table programmes in schools and kura, and healthy food and drink policies in schools and kura. Policies with lower scores included food labelling and reformulation policies and pricing agricultural emissions. The later focus groups and interviews showed contributors generally raised similar ideas regarding the scope, implementation, barriers and unintended consequences for the final six policies. There was consensus that grouping the policies may likely increase their effectiveness, ease implementation, and facilitate strong collaboration between communities and stakeholders in delivering the policies. There was also agreement on centring the policies through a Māori framework and using existing policy examples instead of running pilots, considering the large amount of work already completed by communities to implement these policies without significant government support. Most people regarded parts of pilots such as evaluation as important to identify and correct any issues with policy implementation. The next steps in engagement with policy actors include recognising relevant policies that already exist and looking at which areas of the food system are “entry points” to work on to achieve specific goals to improve food systems.

Policies in educational settings scored highly across different contributor groups and this reflected public support for policy in educational settings generally. The two highest ranked policies: healthy food and drink policies and garden to table programmes in schools and kura are currently implemented in NZ [38–40]. For healthy food and drink policies, the Ministry of Health provides guidance and support to schools to create healthy food and drink policies in schools focused on improving access to healthy foods and drinks through a traffic-light classification system [41]. The most recent review of healthy food and drink policies in schools and kura showed that 38.5% of primary and 44.8% of secondary schools currently had a policy as of 2016 [42]. The Healthy Active Learning programme is a five year programme by Ihi Aotearoa (Sports New Zealand), the Ministry of Health, Te Whatu Ora (Health New Zealand) and the Ministry of Education that supports schools voluntarily in healthy eating, drinking and physical activity [43,44].

Evaluations reflect similar concerns to those raised in the final consultation around implementation, barriers and unintended consequences [43,44]. Identified barriers included resistance from parents and caregivers, unhealthy food environments around schools, and the convenience of processed and ready-to-eat food for students. A project looking at school food environments among primary and secondary schools in Hawke’s Bay showed these barriers to be reported by schools, 14% said there was resistance from parents/whānau, 18% said their healthy food and drink policies were weakened by unhealthy food stores situated near the school [45].

The garden to table programme is delivered by the Garden to Table Trust in NZ, which is a registered charity [39,46]. Schools can register their interest to join the programme at a subsidised cost. Evidence shows that the programme improves teachers’ and students’ food and nutrition knowledge [40]. Current evaluations of the garden to table programme showed that the community links to maintain the gardens during the school holidays were vital to sustaining the programme [40,46]. The long term sustainability of the programme given funding, time and resource challenges was documented in another evaluation where schools reported difficulty obtaining funding after the initial two years of support provided [40]. Additionally, having a dedicated specialist for the kitchen and the garden were vital to maintain the gardens[40]. Equity concerns highlighted by the contributors such as access to land and cost of participation were reflected in a review showing the need for greater certainty and facilitation of food sovereignty and diverse urban foodscapes for urban agriculture in Aotearoa NZ[47]. [48].

A recent evaluation of a community garden in Lyttleton, NZ found key challenges such as land productivity, finances and the capacity of volunteers and staff working in the gardens [49], were all concerns shared by the contributors in the current project. Challenges with land productivity were found as community stakeholders reported conducting extra work to make the land productive, and the original amount of funds for the gardens was insufficient for garden maintenance. Hanna and Pip (2022) outlined how the current policy landscape of legislation, district and regional plan rules and bylaws makes it ’confusing’ and ’complicated’ for local communities [48].

Māra kai, while associated with food production, often also represent a reclamation of food sovereignty by Māori and are a form of resistance to colonial authority especially in relation to land use. [50,51] For example one study investigated Māori women leading local, sustainable food systems and the importance of knowledge transfer [50]. These women discussed the succession of knowledge as exercising tino rangatiratanga (self-determination, leadership, exercise of power) and the importance of passing this knowledge between generations [50].

The evidence on healthy food education shows that public education is effective at addressing learning needs and understanding gaps but it is insufficient as a standalone intervention to address long term population health and nutrition outcomes [51,52]. This finding was reflected by contributors when discussing the need to implement this policy alongside other policies to ensure wider effectiveness. A barrier discussed among contributors was strong commercial opposition. For example, when the United States Dietary Guidelines Advisory Committee (DGAC) recommended that sustainability be incorporated into its dietary guidelines, the recommendations were rejected and remain absent [53,54]. Opposition from the meat industry is considered the most likely reason [54]. Learning how to acknowledge, understand and mitigate potential industry pressures is critical to implementing this policy.

Policies addressing food prices (increasing incomes, removing GST from core food, and making local food cheaper) generally scored well, especially among Māori. An increased focus on food prices is unsurprising given inflation in food commodity prices in NZ, when this study took place [55]. Figures from Statistics New Zealand show that in June 2023 compared with May 2022, food prices increased by 22% for fruit and vegetables, 11% for meat, poultry and fish and 13% for grocery food [55]. An increased awareness of policies to address food prices was reflected in political polling, as a 2023 public poll showed 77% of New Zealanders agreeing that reducing the price of everyday food items should be the top priority for the government [56]. Political parties also included ‘remove GST from food’ in preparation for the 2023 Election in Aotearoa NZ [57], and another poll showed 76% of New Zealanders in support of removing GST from food [19,58].

A 20% fruit and vegetable subsidy has been viewed as more feasible among Aotearoa NZ contributors as other countries do not tax this food group [59]. Evaluations have been conducted on the plausibility of removing GST from food and drink [60,61]. The Tax Working Group report in September 2018 aligned with considerations for implementation, barriers and unintended consequences among the current study’s contributors [61]. The report mentions the barrier of loss of government revenue and unintended consequences of this halting or reducing existing services. Their analysis predicted the policy to benefit higher socioeconomic households more than lower socioeconomic households, with a weekly benefit of $15 for decile 1 (low SES) households compared to $53 for decile 10 households [61]. The working group argues that a more targeted wealth transfer would be more effective. Administration costs were another barrier raised by the working group as the policy requires businesses to find and separate exempted transactions. The overall benefit of the policy to low-income households was not deemed justified compared to these required compliance costs [61].

The greater emphasis on food cost among Māori aligns with the concerns raised by whānau Māori in another study, where the cost of healthy foods was deemed the most significant influence on childhood obesity and food intake [62]. The lived experience of Māori and how policies have failed Māori is reflected in statistics showing that non-Māori are less likely than Māori to be represented in the lowest income bracket (less than $10,000 gross per annum), meaning Māori would benefit most as an ethnic group from income increases [63,64].

As suggested by the contributors in the current study, there is evidence that policies to increase incomes are pro-equity [65–67]. This includes a review of the determinants of food insecurity in Aotearoa NZ concluding that income was the strongest predictor of food insecurity [66]. Given the ethical issues with researchers conducting such studies, there is limited evidence directly assessing the impact of income on healthy and sustainable eating. However, a randomised control trial (RCT) in Aotearoa NZ found that low-income households increased their overall expenditure on food when provided with vouchers to spend on food [67]. Concerns among contributors about whether increasing incomes would also increase the consumption of healthy and sustainable food are plausible, given that no differences between the intervention and control group were found in fruit and vegetable consumption in this RCT. Research has suggested that the UBI could reduce the high levels of health inequities globally, allowing people to maintain or improve healthy living. One study found that expanding an existing food assistance programme (the Supplemental Nutrition Assistance Program or SNAP) to a UBI program could reduce food insecurity by 89% if paid for by those in higher income tax brackets[68].

### Strengths and limitations

Key strengths of this research include the use of policy scoring criteria. These criteria are well supported from previous research and address a range of factors to consider in policy making. The retention rate of contributors was high among those who originally suggested the policies and then scored and discussed these in more detail. For example, the scoring exercise had a 71% retention rate, with only one interviewee unable to attend a follow up interview and all focus group contributors still in their role returned for the focus group. This research is the first known study in Aotearoa NZ to have sustainable kai (food) policies scored and discussed in detail in terms of policy implementation, barriers and intended consequences across a range of contributors from different backgrounds.

There are limitations associated with this research. Policies from engagement with original contributors needed evidence of their effect on dietary intake which would then allow them to be quantitatively modelled. This led to 57% (63) of 111 of policies being excluded. Overall, a bias was seen towards policies with more extensive, published research compared to others, such as new and potentially innovative policies that were cross-sectorial and addressed social and environmental sustainability instead of solely dietary intake. For example, policies that addressed the nutritional value of food such as healthy food policies, food advertising, sugar taxes and food labelling with extensive research dominated the final list of policies included in the policy scoring exercise. Policies such as facilitating land use change and supporting farmers to grow more sustainable food, were excluded. This may also exclude policies published by Indigenous peoples, who are less likely to be published or to seek publication in non-Indigenous academic journals.

## Conclusion

The policy scoring exercise and subsequent focus groups and interviews explored key considerations regarding the scope, implementation, barriers and unintended consequences for six food policies. This research provides rich qualitative data based on the perspectives of a variety of stakeholders. Further research is also needed on policies with weak supporting evidence to add to the political agenda for policymakers. Insights from this research provide clear evidence for policymakers to consider when developing and implementing sustainable dietary and food system policies.

## Data Availability

In keeping with the consent research participants gave, the qualitative and quantitative data this paper is based on is not available to share.

## Acknowledgements

Thank you to our participants for contributing your lived experience and expertise to the project, and continuing to participate in the project. Many thanks to the project advisory group, and the wider project team. This work was supported by the Healthier Lives | He Oranga Hauora National Science Challenge under Grant No. UOOX1902.

## Notes

### Competing Interest Statement

The authors have declared no competing interest.

### Author Declarations

The University of Otago Ethics Committee approved this study on 13th September 2021 (reference no D21/293).

