## Supplementary Table 3 and 4 for "Scoring policies with stakeholders to support healthy, sustainable food (kai) in Aotearoa New Zealand"

### Supplementary material

*Supplementary Table 3: Criteria definitions and policy descriptions*

| **Criteria** | **Definition (statement and example)** |
|---|---|
| How acceptable they would be to a wide range of people | Based on your knowledge, do you think the policy will be acceptable to a wide variety of people (eg. Community members, central and local government, iwi and hapū, non-governmental organisations and the food industry).<br><br>People are likely to agree that the benefits of this policy out-weigh its disadvantages and would agree that it should be implemented in NZ. |
| How possible it would be to implement the policy | How possible it would be to get this policy initiative up and running in New Zealand.<br><br>This policy would be possible to implement in New Zealand considering factors such as cost, infrastructure change and other logistics. |
| Their ability to improve the environment | Based on your knowledge, what is the policies' ability to improve environmental sustainability?<br><br>This policy would promote eating certain foods that have a positive impact on an aspect of environmental sustainability (greenhouse gas emissions, biodiversity, nitrogen use etc.) |
| Their ability to improve diets of New Zealanders | Based on your knowledge, what is the policies' ability to support New Zealanders to eat healthy foods that increase their chances of living a long and healthy life.<br><br>This policy would promote the intake of healthy food which will reduce the risk of diet related diseases such as cancer and heart disease. |
| Its ability to improve the health of Māori | Based on your knowledge, what is the policies' ability to improve health outcomes for Māori and reduce health inequities (differences in health status between Māori and non-Māori).<br><br>This policy would improve Māori health through improving the kai Māori people eat. |

*Supplementary Table 4: Te Reo Māori Glossary*

| Kupu (word) | Definition |
|---|---|
| Aotearoa | Land of the long white cloud, otherwise known as New Zealand (NZ) |
| aroha | Love |
| Atua | Godlike energy, a being of spiritual significance |
| Hākari | Feast of kai |
| Hauora | Health and wellbeing |
| kai | Food, something that gives sustenance |
| kaupapa Māori | A collaborative approach to an issue which is uniquely Māori and is led by Māori for the benefit of Māori. |
| Kaiaranga | Weaver |
| Kupu | Word |
| Kura | School, place of learning. In this paper, kura refers to Māori-language immersion schools. |
| Māori | Person who is Indigenous to Aotearoa and not Moriori |
| māra | Garden, a physical space |
| Māra kai | A place of storytelling, Indigenous praxis and a communal kai resource as well as a place of Rongo-mā-Tane (atua or energy of peace and cultivated foods) and is held by collective interests for collective benefit |
| Marae | The whenua that the people belong to, meet others and which their wharenui lives on. |
| mātauranga | Māori knowledge and scientific praxis |
| Tino rangatiratanga | Absolute authority, autonomy and sovereignty. Self determination and exercise of power. |
| Rongo | Atua (god or energy) of peace and cultivated foods |

| Kupu (word) | Definition |
|---|---|
| tamariki | Children |
| Tangata whenua | The Indigenous people of Aotearoa (excludes Moriori) |
| Tangihanga | Funeral/mourning |
| te taha wairua | The spiritual dimension or aspect of a persons’ health |
| te taha hinengāro | The mental and emotional dimensions of a persons’health |
| Te Tiriti o Waitangi | Treaty of Waitangi the text written in Te Reo Māori |
| tikanga Māori | Māori customary practice, guidance and a set of rules and ways to achieve goals in a culturally safe manner |
| Whānau | family, collective that embraces the individual, biological and/or chosen |
| Whakawhanaungatanga | Process of relationship building through establishing links, usually familial |
| Whenua | Land, placenta |

821
